# Load and fatigue monitoring in instrumental musicians using an online app: A Pilot Study

**DOI:** 10.1101/2022.09.28.22280457

**Authors:** J. Matt McCrary, Sara Ascenso, Paola Savvidou, Séverine Schraft, Lesley McAllister, Emma Redding, Serap Bastepe-Gray, Eckart Altenmüller

## Abstract

**Background/Aims:** High occupational injury rates are reported in musicians, with a career prevalence of up to 89%. Fatigue and playing (over)load are identified as key risk factors for musicians’ injuries. Self-report fatigue management strategies in sport have demonstrated preventive effects. A self-report fatigue management tool for musicians was developed based on a Delphi survey of international experts and hosted in an online app. The aims of this study are to evaluate the content validity and uptake of this new tool, and explore associations between collected performance quality, physical/psychological stress, pain, injury and fatigue data.

**Methods:** University and professional musicians were asked to provide entries into the online app twice per week for one to six months. Entries into the app were designed to take 2-3 minutes to complete and consisted of the following: 6 questions regarding playing load over the previous 72 hours; 5 questions regarding current levels in key physical/psychological stress domains (sleep, recovery, overplaying, pain, fitness); one question self-rating of performance quality over the previous day; one question regarding current musculoskeletal symptoms; a reaction time task to evaluate psychomotor fatigue.

**Results:** N=96 participants provided an average of 2 app entries (range 0 – 43). Increased playing time, rating of perceived exertion (RPE), and feelings of having to ‘play too much’ were consistently associated with increased self-rated performance quality (p≤ .004; 6.7<|t|<2148.5). Increased ratings of feeling fit and recovering well were consistently associated with reduced pain severity (p<.001; 3.8<|t|<20.4). Pain severity was increased (6.5/10 vs. 2.5/10; p<.001) in participants reporting playing-related musculoskeletal disorders (PRMDs; symptoms affecting playing).

**Conclusion:** The prospective value of regular individual self-report playing load, stress, and performance data collection in musicians is clear. However, limited uptake of the online fatigue management app piloted in this study indicates that new approaches to the collection of these data are needed to realize their potential impact.

## Introduction

The extremely high occupational injury rates of musicians are well-documented, with epidemiologic studies noting a career prevalence of playing-related pain and injuries of up to 89%.^1-3^ A substantial portion of musicians captured in these studies also indicated the presence of frequent or permanent painful symptoms (40%^2^) and an injury requiring medical leave in the previous 18 months.^1^ Fatigue and playing (over)load have been identified as key risk factors for a range of injuries in both professional and university student instrumentalists.^1,4,5^

In sport, strategies to manage fatigue have been developed and demonstrated to have preventive benefits.^6-8^ These strategies predominantly utilize consistent monitoring of individual playing/training load (*load = time * intensity of playing/training*) and key indicators of physical/psychological stress to provide a basis for the distinction between normal acute fatigue processes and fatigue and overload states.^9^ From these load and stress data, modifications to training and/or game activities can be judiciously prescribed to reduce injury risk and optimize performance.^6,7^

A 2016 International Olympic Committee consensus statement outlines practical guidelines for such load and stress management strategies:^8^

1. Load must be monitored individually with a daily or weekly frequency for maximum benefit;
2. Both physiological and psychological stressors significantly impact injury risk and must be considered.

A range of monitoring techniques in sport meet the above guidelines, including evaluations of heart rate responses, power output, global positioning system data, and self-report data.^9^

Fatigue management strategies thus provide a promising avenue for pain and injury prevention in musicians. However, the differing environments of athletes and musicians call for the adaptation and validation of fatigue management tools in musical contexts.^10^ While fatigue management programs in sport are aided by structured and/or team environments and more plentiful resources,^9^ the careers and physical demands of musicians are typically more individualized and resources more limited.^11^ Accordingly, based on a Delphi survey of international musicians’ medicine experts,^10^ we developed a low-cost self-report fatigue management tool for musicians hosted in an online app. The aim of this study is to pilot this new fatigue management tool and evaluate its content validity and uptake in university/conservatoire and professional musicians.Secondary aims of this study are to explore relationships between playing-related musculoskeletal disorders (PRMDs), playing load, physical/psychological stress, and practice/performance quality.

## Methods

### Overview and Participants

Study participants were university/conservatoire or full-time professional (i.e. primary income stream related to music performance) musicians who were recruited (*convenience sampling*) from February 2020 – December 2021 through emails and/or in-person presentations to university music schools and conservatoires in Germany, Austria, the United Kingdom, and the United States. Exclusion criteria were the presence of pain or other symptoms that interfered with playing, not being fluent in English or German, and being younger than 18 years of age. All prospective participants enrolled in the study by accessing the online app through a link provided in all recruitment documentation: musiciansfatigue.formr.org. The online app was hosted by formr, a study framework designed to host complex longitudinal studies.^12^

After being informed about study procedures and privacy policies, participants were assigned a study ID number and asked to provide basic demographic information – age; primary instrument; course of study; years playing their primary instrument; height; weight; estimated weekly hours spent playing musical instrument(s). Participants were then instructed to complete the first day’s entry into the fatigue monitoring portion of the app and reminded that entries should be completed bi-weekly for a minimum of one and up to six months. To maximize accessibility, the app was available in English and German language versions and equally functional when accessed from computers and mobile devices. To encourage engagement, participants received weekly study emails containing information relevant to musicians’ health and wellbeing, as well as monthly data reports as applicable. Further, participants completing at least 8 entries (i.e. 4 weeks of bi-weekly entries) were entered into a prize drawing.

### Fatigue Monitoring Tool

The pilot musicians’ fatigue monitoring tool was designed to capture fatigue-related symptoms, music performance quality, playing/practice load, psychomotor performance, and symptoms/illness interfering with playing, comprising the following components:

#### Fatigue-related symptoms

Five key indicators of fatigue-relevant physical and psychological stress identified in a Delphi survey of 28 international musicians’ medicine experts^10^ were assessed using visual analog scales (0-10 scale): *I had pain; I did not get enough sleep; I recovered well physically; I had to play too much; I felt physically fit*. Studies in sport have indicated that self-report physical and psychological stress data are linked to and, in many cases, predictive of fatigue and related performance decrements.^13,14^

#### Music performance quality

Music performance quality was self-assessed for the previous 24 hours using a 0-10 visual analog scale in response to the question ‘*Please rate the overall quality of your musical performances over the past 24 hours’*. Self-ratings of performance quality have been demonstrated to be significantly correlated to performance ratings by outside assessors in prior research.^15^

#### Playing/practice load

Playing and practice load was evaluated for each of the previous three days (*i*.*e. 0-24 hours, 25-48 hours, and 49-72 hours prior to each entry*) using the session rating of perceived exertion (sRPE).^16^ sRPE is a commonly used metric for monitoring training and playing load in sport, and is equal to the product of daily music practice/performance duration (min) and a rating of perceived exertion (RPE) representative of the overall difficulty of the entire playing day (6-20 scale^17^). RPE and self-report diaries have been effectively used to quantify intensity and daily playing duration, respectively, in instrumentalists.^1,18,19^ Further, sRPE has demonstrated retrospective recall reliability for up to three days in sport athletes.^20^

#### Psychomotor performance

Psychomotor performance, assessed using complex reaction time tasks, has been demonstrated to be impaired in fatigue states in sport athletes.^21,22^ The fatigue monitoring tool in the present study included a complex ‘recognition reaction time’ task. This task functioned by presenting participants with three of the first six letters of the alphabet (e.g. a, c, e) as ‘go’ letters and instructing participants to click (computer version) or tap (mobile version) as fast as possible when ‘go’ letters appeared, but not when ‘no go’ letters (e.g. b, d, f) appeared. Twenty trials were completed with each entry into the online app, with the number of correct responses and average reaction time of correct responses recorded. The reaction time task was programmed and hosted in lab.js, which has demonstrated good validity and reliability in accurately capturing reaction time online.^23^

#### Symptoms interfering with playing (PRMD)

The incidence of pain and/or injury interfering with playing over the previous week was assessed by a single yes/no question, utilizing introductory text from the Musculoskeletal Pain Intensity and Interference Questionnaire for Musicians (MPIIQM)(Zaza et al. definition^24^):^25^ ‘*Playing-related musculoskeletal problems are defined as pain, weakness, numbness, tingling, or other symptoms that interfere with your ability to play your instrument at the level to which you are accustomed. This definition does not include mild transient aches and pains. Currently (in the past 7 days) do you have pain/problems that interfere with your ability to play your instrument at the level to which you are accustomed?*’^25^

#### COVID Symptoms

To control for the potentially confounding impacts of COVID symptoms and mandatory quarantine, participants were asked to indicate if they were ‘currently self-isolating due to COVID-19 symptoms, close contacts, or regulations?’ Entries in which participants answered ‘yes’ were excluded from analyses.

### Statistical Analyses

Data for all parameters were categorized as ‘3-day’ – i.e. single app entry – or ‘weekly’ – i.e. average of multiple app entries by the same participant over a 9-day period. Changes in 3-day data were calculated by subtracting values from two app entries occurring within 5 days of each other. Changes in weekly data were calculated by subtracting average weekly values from two consecutive weeks.

Fixed-effects repeated-measures generalized mixed models were used to analyze relationships between three target parameters – self-reported performance quality, pain severity (*i*.*e. ‘I Had Pain’*) ratings, and reaction time (*i*.*e. as an indicator of psychomotor fatigue*) – and all other investigated parameters. Links between 3-day and weekly data (‘Values’) and 3-day and weekly change data (‘Change’) from target and all other parameters were analyzed. Additionally, potential predictive relationships between 3-day and weekly Change data and Values of the corresponding time-period were analyzed. The Satterthwaite approximation^26^ was used to account for unevenly distributed data, linear and non-linear models were used as appropriate to analyze normal and non-normally distributed target parameter data, and the Bonferroni-Holm correction was used to account for multiple comparisons.^27^ Additionally, exploratory independent samples t-tests, with the Bonferroni-Holm correction, were used to analyze differences in parameters when a PRMD was vs. was not reported in the same data entry. All statistical analyses were performed in SPSS v.27.0 (IBM, Armonk, NY).

### Study Registration and Ethics Approval

This study was prospectively registered in the Australian New Zealand Clinical Trials Registry (ACTRN12619001108101) and approved by the Leibniz University Hannover Central Ethics Committee (EV LUH 12/2019) and Conservatoires UK Ethics Committee (CUK/TL/2019/20/9).

## Results

### Participants and Data Entries

N= 96 participants (N=86 university/conservatoire students; N=10 full-time professionals) enrolled in the study, providing a total of 478 data entries (median app entries per participant = 2; maximum participant entries = 43; minimum participant entries = 0). N=14 participants (N=12 students; N=2 professionals) provided informed consent and enrolled in the study but did not complete any valid data entries. Valid reaction time data were available for 417 data entries. Student participants were from universities in the United States (*N=31*), Germany (*N=29*), the United Kingdom (*N=22*), Austria (*N=2*), Canada (*N=1*), and Ireland (*N=1*). Participants practiced and played for, on average, 480 minutes (*SD = 365; minimum = 0; maximum = 2160*) over the preceding 3-day period, with an average RPE of 12 (*SD = 2; minimum = 6; maximum = 17*). See Table 1 for participant demographics.

**Table 1.** Participant (N=96) Demographics.

| ***Table 1. Participant (N=96) Demographics*** |  |  |
| --- | --- | --- |
| **Mean age** (*Standard deviation; range*) |  | 24 (6; 18 – 55) |
| **Full-time students : Full-time professionals** |  | 86 : 10 |
| **Mean years music performance experience** (*Standard deviation; range*) |  | 14 (7; 4 – 47) |
| **Gender** |  |  |
|  | Male | 38 |
|  | Female | 58 |
| **Instrument** |  |  |
|  | Keyboard | 25 |
|  | Upper strings | 23 |
|  | Woodwinds | 16 |
|  | Brass | 8 |
|  | Vocalists | 8 |
|  | Lower strings | 5 |
|  | Percussion | 5 |
|  | Harp | 3 |
|  | Accordion | 2 |
|  | Guitar | 1 |

### Correlates of Performance Quality (Table 2)

Increased playing time, RPE, and feelings of having to ‘play too much’ were consistently associated with increased self-rated performance quality across 3-day and weekly comparisons (p≤ .004; 6.7<|t|<2148.5). Further, Changes (*increases*) in RPE and feelings of having to ‘Play too much’ over the prior 3 days and week were both associated with higher self-rated performance quality (p≤ .001; 4.3<|t|<10.2). Reaction time, playing load, and ratings of feeling fit, recovering well, not getting enough sleep, and pain severity were also less consistently associated with self-rated performance quality (p<.01; 2.6<|t|<2217.6).

**Table 2.** Associations between self-rated performance quality and all other parameters. Value vs. Value and Change vs. Change comparisons display concurrent relationships. Value vs. Change comparisons represent predictive relationships. Statistically significant associations are highlighted in bold and grey.

| **Performance Quality Rating** | **3-Day Value** |  | **Weekly Value** |  | **3-Day Change** |  | **Weekly Change** |  |
| --- | --- | --- | --- | --- | --- | --- | --- | --- |
| *Parameter* | *Coeff.* | *p* | *Coeff.* | *p* | *Coeff.* | *p* | *Coeff.* | *p* |
| Not enough sleep (value) | .001 | .975 | .042 | .329 |  |  |  |  |
| Not enough sleep (change) | -.028 | .441 | .020 | .744 | -.021 | .466 | **.093** | **.002** |
| Play too much (value) | **.192** | **.004** | **.21** | **<.001** |  |  |  |  |
| Play too much (change) | **.155** | **<.001** | **.304** | **<.001** | **.165** | **<.001** | **.237** | **<.001** |
| Felt fit (value) | **.205** | **<.001** | **.400** | **<.001** |  |  |  |  |
| Felt fit (change) | -.024 | .659 | .157 | .102 | .021 | .664 | **.142** | **.009** |
| Recovered well (value) | **.171** | **.001** | **.286** | **<.001** |  |  |  |  |
| Recovered well (change) | -.119 | .032 | -.169 | .098 | -.013 | .557 | .018 | .229 |
| Pain 0-10 rating (value) | -.056 | .535 | **-.122** | **<.001** |  |  |  |  |
| Pain 0-10 rating (change) | .069 | .123 | .040 | .718 | .023 | .656 | <.001 | .98 |
| Playing load (value) | <.001 | .016 | **<.001** | **<.001** |  |  |  |  |
| Playing load (change) | **<.001** | **.002** | <.001 | .041 | **<.001** | **<.001** | **<.001** | **<.001** |
| Playing time (value) | **.001** | **.001** | **<.001** | **<.001** |  |  |  |  |
| Playing time (change) | **.001** | **<.001** | <.001 | .036 | **.003** | **<.001** | **.001** | **<.001** |
| Rating of perceived exertion (RPE)(value) | **.368** | **<.001** | **.520** | **<.001** |  |  |  |  |
| Rating of perceived exertion (RPE)(change) | **.197** | **<.001** | **.432** | **<.001** | **.292** | **<.001** | **.526** | **<.001** |
| Average reaction time (value) | -.003 | .026 | -.003 | .017 |  |  |  |  |
| Average reaction time (change) | .001 | .664 | **<.001** | **<.001** | <.001 | .783 | <.001 | .991 |
| Reaction time, number correct (value) | -.012 | .792 | .102 | .181 |  |  |  |  |
| Reaction time, number correct (change) | .034 | .564 | -.097 | .475 | .079 | .262 | .063 | .268 |
| Reaction time, time*correct (value) | <.001 | .041 | <.001 | .130 |  |  |  |  |
| Reaction time, time*correct (change) | **<.001** | **.001** | .002 | .431 | **.001** | **<.001** | **<.001** | **.004** |

### Relationships with pain severity (Table 3)

Increased ratings of feeling fit and recovering well were consistently associated with reduced pain severity (p<.001; 3.8<|t|<20.4). All other investigated parameters were less consistently associated with pain severity ratings (p<.001; 3.5<|t|<913.4). No changes in any parameter were consistently linked to pain severity values.

**Table 3.** Associations between pain severity (*‘I Had Pain’*) ratings and all other parameters. Value vs. Value and Change vs. Change comparisons display concurrent relationships. Value vs. Change comparisons represent predictive relationships. Statistically significant associations are highlighted in bold and grey.

| **Pain Severity Rating** | **3-Day Value** |  | **Weekly Value** |  | **3-Day Change** |  | **Weekly Change** |  |
| --- | --- | --- | --- | --- | --- | --- | --- | --- |
| *Parameter* | *Coeff.* | *p* | *Coeff.* | *p* | *Coeff.* | *p* | *Coeff.* | *p* |
| Not enough sleep (value) | -.119 | .006 | **-.667** | **<.001** |  |  |  |  |
| Not enough sleep (change) | .055 | .233 | -.237 | .007 | **.151** | **<.001** | **.170** | **<.001** |
| Play too much (value) | **-.388** | **<.001** | **-.278** | **<.001** |  |  |  |  |
| Play too much (change) | .233 | <.001 | **-.457** | **<.001** | <.001 | .988 | .080 | .029 |
| Felt fit (value) | **-.524** | **<.001** | **-.808** | **<.001** |  |  |  |  |
| Felt fit (change) | -.018 | .814 | <.001 | .998 | **-.178** | **<.001** | **-.223** | **<.001** |
| Recovered well (value) | **-.529** | **<.001** | **-.715** | **<.001** |  |  |  |  |
| Recovered well (change) | -.127 | .052 | .129 | .413 | **-.246** | **<.001** | **-.399** | **<.001** |
| Performance quality rating (value) | **-.868** | **<.001** | **-.625** | **<.001** |  |  |  |  |
| Performance quality rating (change) | -.027 | .661 | -.026 | .818 | .023 | .656 | <.001 | .989 |
| Playing load (value) | **>-.001** | **<.001** | **>-.001** | **<.001** |  |  |  |  |
| Playing load (change) | >-.001 | .519 | -.002 | .170 | <.001 | .983 | **<.001** | **<.001** |
| Playing time (value) | **-.001** | **<.001** | -.002 | .318 |  |  |  |  |
| Playing time (change) | >-.001 | .271 | <.001 | .998 | <.001 | .986 | **<.001** | **.001** |
| Rating of perceived exertion (RPE)(value) | **-.431** | **<.001** | **-.423** | **<.001** |  |  |  |  |
| Rating of perceived exertion (RPE)(change) | .006 | .915 | -.127 | .248 | <.001 | .993 | **.158** | **<.001** |
| Average reaction time (value) | <.001 | .788 | .007 | .007 |  |  |  |  |
| Average reaction time (change) | **-.003** | **<.001** | -.009 | .017 | <.001 | .999 | .001 | .078 |
| Reaction time, number correct (value) | -.223 | .036 | **-1.00** | **<.001** |  |  |  |  |
| Reaction time, number correct (change) | .116 | .237 | .080 | .721 | <.001 | .995 | .059 | .058 |
| Reaction time, time*correct (value) | **>-.001** | **<.001** | **<.001** | **<.001** |  |  |  |  |
| Reaction time, time*correct (change) | .001 | .145 | **-.021** | **<.001** | <.001 | .941 | <.001 | .125 |

### Relationships with reaction time (psychomotor speed) (Table 4)

No parameters were consistently associated with reaction time. Further, no changes in any parameter over the previous 3 days or week were significantly associated with reaction time (p>.004; |t|<3.1). Playing load, playing time, RPE, and ratings of feeling fit, recovering well, pain severity, and performance quality were intermittently associated with reaction time (p<.002; 3.2<|t|<10.6).

**Table 4.** Associations between reaction time and all other parameters. Value vs. Value and Change vs. Change comparisons display concurrent relationships. Value vs. Change comparisons represent predictive relationships. Statistically significant associations are highlighted in bold and grey.

| **Reaction Time** | **3-Day Value** |  | **Weekly Value** |  | **3-Day Change** |  | **Weekly Change** |  |
| --- | --- | --- | --- | --- | --- | --- | --- | --- |
| *Parameter* | *Coeff.* | *p* | *Coeff.* | *p* | *Coeff.* | *p* | *Coeff.* | *p* |
| Not enough sleep (value) | -.378 | .763 | -1.357 | .368 |  |  |  |  |
| Not enough sleep (change) | 1.316 | .308 | -.742 | .726 | .304 | .005 | -.397 | .702 |
| Play too much (value) | -1.640 | .159 | -1.821 | .101 |  |  |  |  |
| Play too much (change) | -.161 | .925 | -5.686 | .037 | .927 | .559 | 2.661 | .041 |
| Felt fit (value) | **-5.911** | **<.001** | -3.850 | .034 |  |  |  |  |
| Felt fit (change) | -1.016 | .602 | -.927 | .799 | -2.269 | .228 | -1.735 | .433 |
| Recovered well (value) | **-6.197** | **<.001** | **-8.400** | **<.001** |  |  |  |  |
| Recovered well (change) | -.984 | .555 | 8.045 | .015 | -4.003 | .025 | .643 | .713 |
| Pain 0-10 rating (value) | -.077 | .952 | **4.699** | **.001** |  |  |  |  |
| Pain 0-10 rating (change) | .496 | .814 | 3.898 | .354 | 2.195 | .278 | 2.303 | .23 |
| Performance quality rating (value) | -3.85 | .034 | **-7.149** | **<.001** |  |  |  |  |
| Performance quality rating (change) | 1.736 | .364 | -1.493 | .315 | -3.932 | .027 | .972 | .548 |
| Playing load (value) | **-.002** | **.001** | **-.002** | **<.001** |  |  |  |  |
| Playing load (change) | -.001 | .235 | -.002 | .006 | -.002 | .085 | -.001 | .056 |
| Playing time (value) | **-.030** | **.001** | **-.029** | **<.001** |  |  |  |  |
| Playing time (change) | -.017 | .151 | -.029 | .004 | -.025 | .096 | -.007 | .183 |
| Rating of perceived exertion (RPE)(value) | -3.264 | .009 | **-5.333** | **<.001** |  |  |  |  |
| Rating of perceived exertion (RPE)(change) | .601 | .664 | -3.665 | .108 | -1.670 | .241 | **-3.289** | **<.001** |
| Reaction time, number correct (value) | **-5.89** | **.001** | **-17.308** | **<.001** |  |  |  |  |
| Reaction time, number correct (change) | 6.111 | .015 | 9.948 | .011 | -3.015 | .272 | 1.291 | .636 |

### Symptoms interfering with playing (PRMD)

Symptoms interfering with playing (PRMDs) were reported by 23 participants (18 students/5 professionals; Primary instruments: piano (4), violin (3), vocal (3), accordion (2), bass (2), saxophone (2), viola (2), flute (1), guitar (1), horn (1), clarinet (1), oboe (1)) across 32 of the total 478 (6.7%) data entries. Pain severity was significantly greater when symptoms interfering with playing were reported (|t|=8.9; p<.001)(Figure 1). Additionally, several parameters trended towards significance in PRMD vs. No PRMD comparisons: increased 3-day RPE (PRMD (*mean ± standard deviation*): 12.6±2.3; No PRMD: 11.5±2.3; p=.02) and feelings of playing too much (p=.049; Figure 1); and decreased feelings of fitness (p=.03; Figure 1) and recovering well (p=.02; Figure 1). 3-day playing time (PRMD: 473.1±363.6 minutes; No PRMD: 577.7±379.3 minutes), 3-day playing load (PRMD: 6168.8±4997.9; No PRMD: 7934.3±5845.3), and reaction time (PRMD: 643.8±88.7 milliseconds; No PRMD: 674.7±92.5 milliseconds) did not significantly differ when PRMDs were vs. were not reported (p>.07).

**Figure 1.**
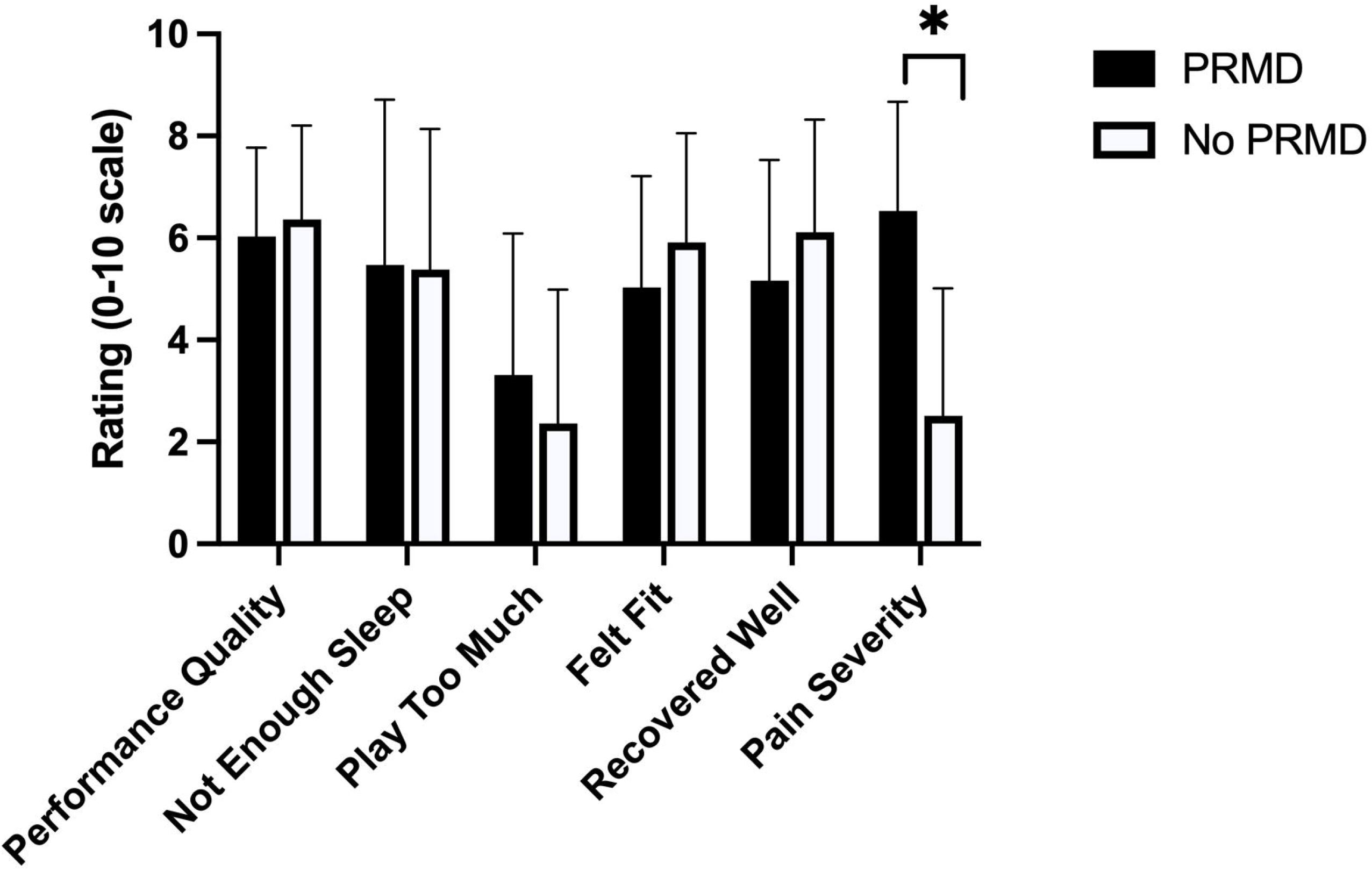
Self-report performance and physical/psychological stress outcomes with (PRMD) and without (No PRMD) concurrent reports of symptoms affecting playing (PRMD). Data presented as mean ± standard deviation. * - significant difference after Bonferroni-Holm correction for multiple comparisons; p < .001.

## Discussion

This study demonstrates the potential of regularly collected self-report playing load and physical/psychological stress data to advance understanding of the complex influences of these parameters on performance, pain and injury outcomes in musicians. However, limited uptake of the piloted fatigue management tool diminishes its prospective impact and raises questions regarding the feasibility of regular self-report data collection in high-level musicians.

Unexpectedly, increased RPE and feelings of having to ‘play too much’ were strongly and consistently associated with *increased* self-rated performance quality. Increased playing time and playing load were also linked to *increased* self-rated performance quality, albeit slightly less consistently, in line with prior study.^15^ Self-rated music performance quality has been shown to be significantly correlated with performance quality ratings of external assessors.^15^ However, the only moderate strength of this prior association favors discussion focusing on associations between investigated parameters and *perceived* performance quality.

Links between increased performance quality and feelings of having to ‘play too much’ indicate that musicians may benefit from a version of the post-activation potentiation response observed in athletes, where prior fatiguing activity leads to short-term gains in performance.^28^ Links between increased RPE and improved self-rated performance mirror prior study associating effort and performance outcomes.^29^ Increased RPE and playing load have been assumed to be a negative outcome for both performance and injury risk. Accordingly, this study adds to a growing body of evidence challenging assumptions that increased effort and playing load have a universally negative impact on injury and performance quality,^19,30^ instead suggesting more complex interactions between playing load, exertion, injury, and performance.^31^ A small sample of reports of PRMDs in the present study precludes conclusive comments regarding the links between playing load and injury/PRMD incidence.

Associations between increased feelings of physical fitness and feelings of recovering well physically and reduced pain severity are consistent with prior studies demonstrating links between increased physical fitness and reduced pain severity.^30,32^ Study participants also indicated that low levels of pain (mean 2.5/10) did not interfere with playing; this result is, once again, consistent with prior research asserting that low-level pain symptoms are common in musicians and do not necessarily impact playing.^33^ Significantly increased pain intensity (mean 6.5/10) was associated with reports of PRMDs, indicating that one benefit of increased physical fitness in musicians may be reported analgesic effects which may globally reduce pain severity to manageable levels without pathological injury.^34^ Further research into relationships between fitness, pain and PRMDs in musicians is required to confirm this hypothesis.

The absence of consistent associations of reaction time, an indicator of psychomotor fatigue in athletes,^21,22^ with playing load or physical/psychological stress outcomes in the present study has multiple explanations. This result could be explained by differential fatigue processes in musicians vs. athletes,^35^ but also by low numbers of consecutive data entries which limit statistical power. Additional study is needed to determine whether reaction time is a valid indicator of fatigue states in musicians.

Analyses of associations between study outcomes, as well as assessment of the content validity and prospective utility of the pilot fatigue monitoring tool, are ultimately limited, however, by challenges with uptake in professional and student musicians. Despite a recruitment push across multiple large international music universities and conservatoires and multiple incentives for participation, only 96 participants were enrolled in 22 months and the average participant engaged with the fatigue management tool for just 1 week. While app pre-testing indicated good functionality, the unvalidated pilot app may not have been conceptually appealing and/or presented enticingly enough for musicians with many competing priorities. Recruitment difficulties were also likely exacerbated by the conduct of the study during various stages of COVID lockdowns and remote university/conservatoire learning from 2020-21 – multiple authors noted substantial impacts of the pandemic on the engagement of their students. Further, our app relied on participation prompted by email reminders, with analyses of data explicitly not provided to study participants to avoid influencing practice and performance behaviors. Fatigue monitoring data in sport are typically collected and analyzed by staff on an ongoing basis,^9^ which likely increases long-term engagement. Further research using ongoing staff data collection/analysis methods in musicians is needed to determine its impact on both engagement and observed results.

Alternately and/or additionally, recruitment difficulties could underscore the importance of ongoing international work to enhance musicians’ health literacy.^36^ Low health literacy in musicians is hypothesized to present a critical barrier to engagement of musicians in health-promoting practices. Mandatory health education seminars and coursework are integrated into the curricula of an increasing number of conservatoires and university music programs, including the majority of collaborating institutions in this study. However, such integrated health education may not yet translate into general enthusiasm for new approaches to health promotion and enhancement such as the novel app presented in this study. Further research into motivations influencing the uptake of novel approaches to health promotion in musicians is needed to provide further insights and enhance recruitment practices in future studies.

In conclusion, this study demonstrates the prospective value of regular individual self-report playing load, stress, and performance data collection in musicians. However, limited uptake of the online fatigue management app piloted in this study indicates that new approaches to the collection of these data are needed to realize their potential impact.

## Data Availability

All data produced in the present study are available upon reasonable request to the authors

## Acknowledgements

J. Matt McCrary was supported by a Postdoctoral Fellowship from the Alexander von Humboldt Foundation.

## Notes

### Competing Interest Statement

The authors have declared no competing interest.

### Clinical Trial

ACTRN12619001108101

### Funding Statement

This study was supported by a Postdoctoral Fellowship from the Alexander von Humboldt Foundation (to J. Matt McCrary).

### Author Declarations

The Central Ethics Committee of Leibniz University Hannover gave ethical approval for this work. The Ethics Committee of Conservatoires UK also gave ethical approval for this work (CUK/TL/2019/20/9).

